# Rho GTPase Activating Protein 15 (ARHGAP15) increases cancer risk in patients with diverticular disease

**DOI:** 10.1101/2022.10.30.22281714

**Authors:** Steven Lehrer, Peter H. Rheinstein

## Abstract

**Introduction:** Patients with diverticular disease who have colorectal histopathology, normal or abnormal, have increased risk of overall incident cancer. To determine whether the cancer risk might have a genetic basis, we performed a Genome Wide Association Study (GWAS) with data from UK Biobank.

**Methods:** We used PLINK 2, a whole-genome association analysis toolset, to analyze the UKB chromosome files. To identify cases, we used ICD10 code K57.3, diverticular disease of large intestine without perforation or abscess. We selected subjects younger than 45, since genetic forms of disease tend to manifest in younger patients.

**Results:** GWAS summary (Manhattan) plot of the meta-analysis association statistics, highlighting one susceptibility locus, Rho GTPase Activating Protein 15 (ARHGAP15), with genome wide significance for diverticulosis.

**Conclusion:** We conclude that ARHGAP15 could be responsible for the association of diverticular disease and cancer.

---

Ma et al studied cancer risk in diverticular disease and reported that patients with diverticular disease who have colorectal histopathology, normal or abnormal, have increased risk of overall incident cancer [1]. The cancers included colorectal cancer, lung cancer, pancreatic cancer, and liver cancer. To determine whether the cancer risk might have a genetic basis, we performed a Genome Wide Association Study (GWAS) with data from UK Biobank [2].

## Methods

We used PLINK 2, a whole-genome association analysis toolset, to analyze the UKB chromosome files. We adhered to recommended quality control procedures [3] that consisted of the following:

1. Missingness of SNPS 0.05: This command excluded SNPs that are missing in a large proportion of the subjects. In this step, SNPs with low genotype calls were removed.
2. Missingness of individuals 0.05: This command excluded individuals who had high rates of genotype missingness. In this step, individuals with low genotype calls were removed.
3. Hardy Weinberg equilibrium 1e-6: This command excluded markers which deviate from Hardy–Weinberg equilibrium.
4. Minor allele frequency (MAF) threshold 0.01: This command included only SNPs above the set MAF threshold.

To identify cases, we used ICD10 code K57.3, diverticular disease of large intestine without perforation or abscess. We selected subjects younger than 45, since genetic forms of disease tend to manifest in younger patients [4]. We analyzed data from 1828 subjects, 50.5% women and 49.5% men.

## Results

GWAS summary (Manhattan) plot of the meta-analysis association statistics, highlighting one susceptibility locus, Rho GTPase Activating Protein 15 (ARHGAP15), with genome wide significance for diverticulosis, is shown in figure 1A. The red line indicates the genome wide significance threshold of a P value less than 5×10^−8^.

**Figure 1.**
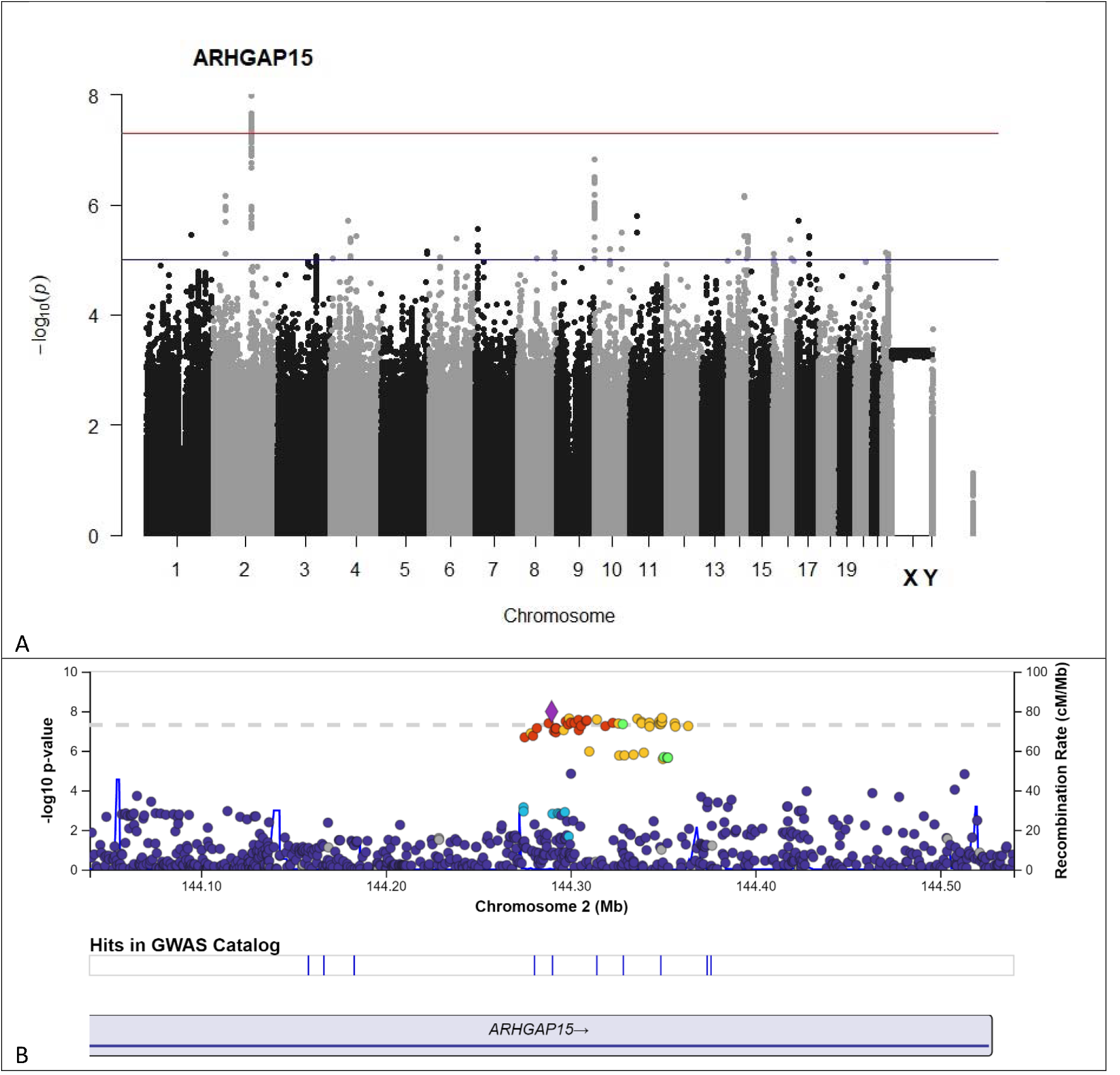
(A) GWAS Summary (Manhattan) Plot of the meta-analysis association statistics highlighting one susceptibility locus (ARHGAP15) on chromosome 2q22 with genome wide significance for colon diverticula. The red line indicates the genome wide significance threshold of a p value less than 5×10^−8^. (B) LocusZoom plot of ARHGAP15 association. Genomic position is depicted on the x-axis. The left y-axis shows the –log10 of the p-value. SNPs are colored based on their correlation (r^2^) with the labeled top SNP, rs7607879 (purple diamond), which has the smallest p value in the region. The fine-scale recombination rates estimated from 1000 Genomes (EUR) data (right y axis) are indicated by the fluctuating blue line. The position of ARHGAP15 relative to rs7607879 is displayed.

LocusZoom plot of ARHGAP15 association is in figure 1B. Genomic position is depicted on the x-axis. The left y-axis shows the –log10 of the p-value. SNPs are colored based on their correlation (r^2^) with the labeled top SNP, rs7607879 (purple diamond), which has the smallest p value in the region. The fine-scale recombination rates estimated from 1000 Genomes (EUR) data (right y axis) are indicated by the fluctuating blue line. The position of ARHGAP15 relative to rs7607879 is displayed.

The SNP rs7607879 is an intron variant, position chr2:143532194, alleles C>T, reference allele (C) frequency 0.2.

## Discussion

ARHGAP15 inhibits PAK kinase. The regulation of cell motility, survival, metabolism, cell cycle, proliferation, transformation, stress, inflammation, and gene expression are just a few of the critical activities that PAK kinases affect. PAK kinase dysregulation interferes with cellular homeostasis and negatively affects essential cell activities, many of which are linked to a variety of human disorders, including cancer.

ARHGAP15 is associated with diverticular disease and expressed in the colon [5]. ARHGAP15 may be a tumor suppressor during colorectal cancer progression [6] and regulates lung cancer cell proliferation and metastasis via the STAT3 pathway [7].

We conclude that ARHGAP15 could be responsible for the association of diverticular disease and cancer that Ma et al reported [1].

## Data Availability

data available from UK Biobank with approved project

https://www.ukbiobank.ac.uk/

## References

1. Ma W, Walker MM, Thuresson M, et al. Cancer risk in patients with diverticular disease: a nationwide cohort study. J Natl Cancer Inst 2022; 10.1093/jnci/djac190.

2. Arthur RS, Wang T, Xue X, et al. Genetic Factors, Adherence to Healthy Lifestyle Behavior, and Risk of Invasive Breast Cancer Among Women in the UK Biobank. J Natl Cancer Inst 2020;112(9):893–901.

3. Marees AT, de Kluiver H, Stringer S, et al. A tutorial on conducting genome-wide association studies: Quality control and statistical analysis. International journal of methods in psychiatric research 2018;27(2):e1608.

4. Jiang X, Holmes C, McVean G. The impact of age on genetic risk for common diseases. PLoS Genet 2021;17(8):e1009723.

5. Sigurdsson S, Alexandersson KF, Sulem P, et al. Sequence variants in ARHGAP15, COLQ and FAM155A associate with diverticular disease and diverticulitis. Nat Commun 2017;8:15789.

6. Pan S, Deng Y, Fu J, et al. Decreased expression of ARHGAP15 promotes the development of colorectal cancer through PTEN/AKT/FOXO1 axis. Cell Death Dis 2018;9(6):673.

7. Liu ZD, Mou ZX, Che XH, et al. ARHGAP15 regulates lung cancer cell proliferation and metastasis via the STAT3 pathway. Eur Rev Med Pharmacol Sci 2019;23(13):5840–5850.

